# Preventable deaths involving falls in England and Wales, 2013-2022: a systematic case series of coroners’ reports

**DOI:** 10.1101/2023.05.27.23290640

**Authors:** Kaiyang Song, Clara Portwood, Jessy Jindal, David Launer, Harrison S France, Molly Hey, Georgia Richards, Francesco Dernie

**Affiliations:** Medical Sciences Division, University of Oxford, Oxford, OX3 9DU, UK; Centre for Evidence-Based Medicine, Nuffield Department of Primary Care Health Sciences, University of Oxford, Radcliffe Observatory Quarter, Woodstock Road, Oxford, OX2 6GG, UK; Oxford University Hospitals NHS Trust, Oxford, OX3 9DU, UK

## Abstract

**Background:** Falls in older people are common, but can lead to significant harm including death. Coroners in England and Wales have a duty to report cases where action should be taken to prevent deaths, but dissemination of their findings remains poor.

**Objective:** To identify preventable fall-related deaths, classify coroners’ concerns, and explore organisational responses.

**Design:** Retrospective case series.

**Setting:** Coroners’ reports to Prevent Future Deaths (PFD) in England and Wales.

**Methods:** Web scraping was used to screen and read PFDs from the Courts and Tribunals Judiciary website from July 2013 (inception) to November 2022. Demographic information, coroners’ concerns and responses from organisations were extracted. Descriptive statistics and content analysis were used to synthesise data.

**Results:** 527 PFDs (12.5% of all PFDs) involved a fall that contributed to death. These deaths predominantly affected older people (median 82 years) in the community (72%), with subsequent death in hospital (70.8%). A high proportion of cases experienced fractures, major bleeding or head injury. Coroners frequently raised concerns regarding falls risks assessments, failures in communication, and documentation issues. Only 56.7% of PFDs received a response from the intended recipients. Organisations produced new protocols, improved training, and commenced audits in response to PFDs.

**Conclusions:** One in eight preventable deaths reported in England and Wales involved a fall. Addressing concerns raised by coroners should improve falls prevention and care following falls especially for older adults. Poor responses to coroners may indicate that actions are not being taken. Wider learning from PFD findings may help reduce preventable fall-related deaths.

## Introduction

Falls are the second largest cause of unintentional injury deaths worldwide [1]. In 2020, there were 6,410 deaths reported from accidental falls in England and Wales, with 5,850 of these affecting persons ≥65 years old [2]. Falls can lead to traumatic injury, such as hip fracture [3] or head injury, which may lead directly to mortality. Additionally, falls can act as the initiating factor in a chain of events that lead to death, such as immobility after a fall precipitating a fatal pneumonia [4].

Preventing significant morbidity and mortality related to falls remains a challenge both in inpatient, care home, and other community settings. Inpatient falls are the most reported patient safety incident in England and Wales [5], while falls in the community led to 223,101 emergency hospital admissions in people aged ≥65 during 2021-2022 [6]. Consequently, preventing falls in hospitals and the community is a national priority as highlighted in the NHS Patient Safety Strategy [7], the NHS Long Term Plan [8], and an essential research need [9]. Although not all falls are preventable, it is important to understand what factors preceding and following a fall are modifiable to prevent serious harm.

Since 1984, coroners have had a duty to report and communicate a death where the coroner believes that action should be taken to prevent future deaths [10]. These reports, named Prevention of Future Deaths (PFDs), are mandated under Paragraph 7 of Schedule 5, Coroners and Justice Act 2009, and regulations 28 and 29 of The Coroners (Investigations) Regulations 2013 [11, 12]. These reports are sent to adressees who must then respond within 56 days. PFDs are now cited as an official source of data for the NHS Patient Safety Strategy [7]. However, there is no mechanism for systematically synthesizing PFDs to identify and disseminate trends and learnings from coroners’ concerns and actions taken or proposed in their responses.

A detailed analysis of PFDs implicating falls has not yet been conducted, which could highlight important learnings for clinical practice and policy to prevent falls and reduce the risk of death following a fall. The aim of our study was to conduct a systematic case series of PFDs to identify and characterise deaths involving falls, synthesise coroners’ concerns, and explore the responses of individuals or organisations to whom PFDs were addressed.

## Methods

A systematic case series was designed, and the study protocol was preregistered on an open repository [13].

### Data collection, screening, and eligibility

Data were acquired from the Courts and Tribunals Judiciary website [14] on 16^th^ November 2022 using a computer programme called a web scraper designed by a study author (FD) and made openly available [15], similar to the Preventable Deaths Tracker (https://preventabledeathstracker.net/). The web scraper automatically downloaded all published PFD portable document formats (pdfs) from the Judiciary website and searched for pre-determined keywords. The keywords chosen were ‘fall’, ‘falls’, ‘falling’, ‘fallen’, ‘fell’ and the positive control word ‘coroner’ (to identify documents which couldn’t be automatically screened for the keywords). The code identified and downloaded 4305 pdfs, of which 4205 were PFDs and the remainder were mis-labelled response letters. Nearly half (48%; n=2010) of the pdfs were readable by code and eligible for primary inclusion and were screened by at least one author (KS, FD) for validation purposes (Supplementary Appendix Figure 1). The remaining 2295 pdfs were independently screened by at least two study authors (KS, CP, JJ, DL, HF, MH). Cases were included if a fall was believed to have caused or contributed to death. Cases were excluded if the fall was precipitated by violence (e.g., a push by another care home resident), or if suicidal intent was suspected by coroners. Any ambiguities regarding inclusion of cases were discussed with and resolved by the senior investigator (FD).

**Figure 1.**
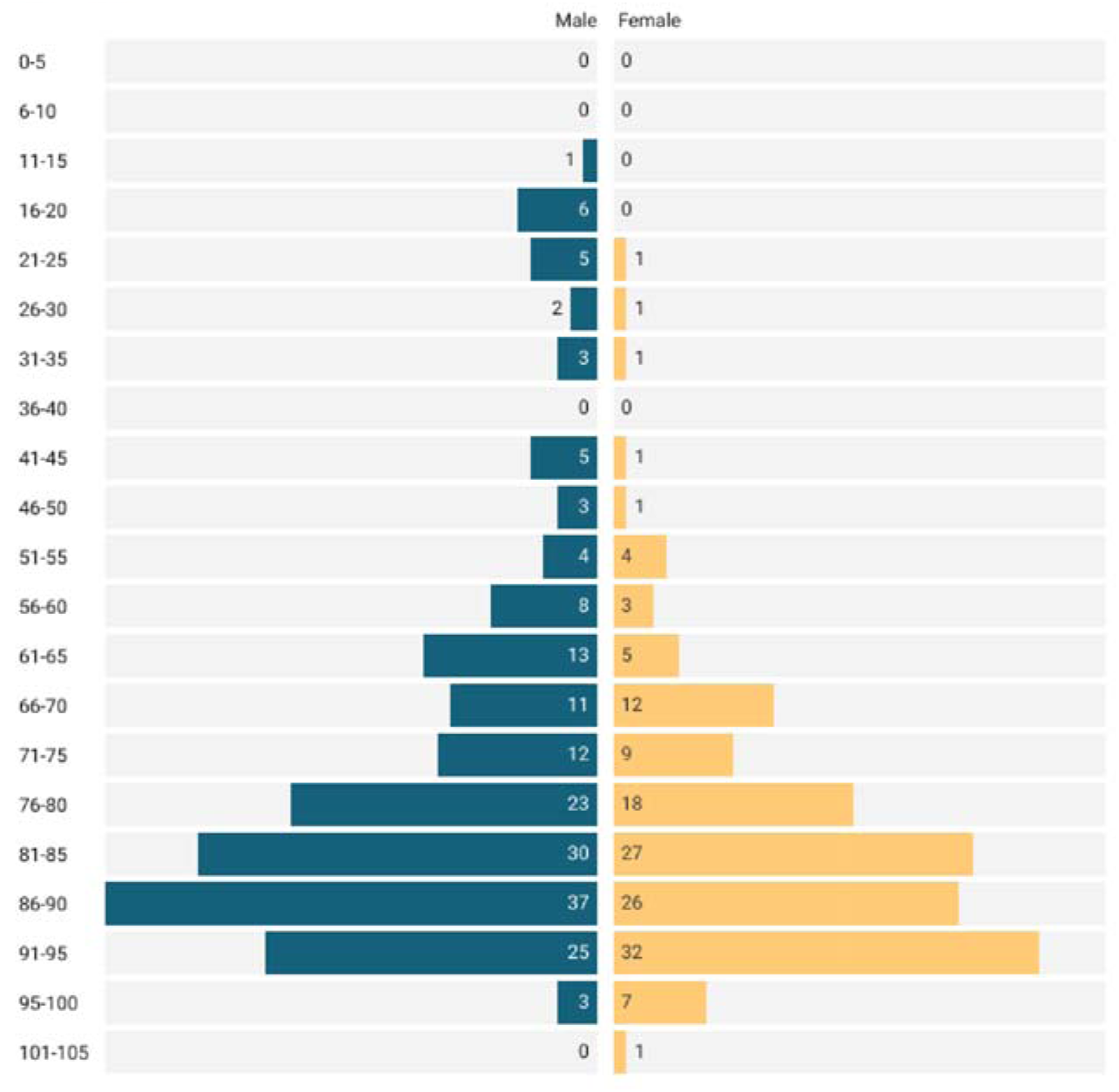
Age distribution of included fall-related PFD cases, by sex. 21.1% of falls and 70.8% of deaths occurred in hospitals. 70.0% of falls occurred in community settings; 30.6% occurred in care home/supported living settings, 25.8% in patients’ own homes, and 15.6% in other community settings (Figure 2, Supplementary Appendix Table 2).

### Data extraction

Case demographics, causes of deaths and morbidity, fall risk factors highlighted by national falls prevention authorities [3, 5], coroners’ concerns, and responses from recipients receiving PFDs were manually extracted by authors (KS, CP), and reviewed (FD). Data regarding the geographic distribution of all PFDs was extracted from the Preventable Deaths Tracker [16].

### Data analysis

The number of included fall-related PFDs, their rates as a proportion of all PFDs, and ONS mortality data were calculated over time. Medians and interquartile ranges (IQRs) were calculated for continuous variables (e.g., age) and frequencies were reported for categorical variables (e.g., sex, location of fall, location of death, coroner area of jurisdiction).

We calculated the years of life lost (YLL) [17] for each case (where age was reported) by extracting their remaining life expectancy from the Office for Nation Statistics (ONS) cohort life tables [18].

Two investigators (CP, KS) assigned the International Statistical Classification of Disease and Related Health Problems 11^th^ Revision (ICD-11) [19] numeric codes for the causes of death to each PFD.

To examine geographical variation, we graphed both the absolute counts and rates of fall-related PFDs per all PFDs in each coroner area mapped to the standard regions of England and Wales.

To evaluate reporting of fall-related risk factors reported in PFDs, information on risk factors detailed in the National Audit of Inpatient Falls (NAIF) [3] and Office for Health Improvement and Disparities (OHID) [5] were collated reported as a percentage of total cases. The NAIF has identified the following risk factors for falls: visual impairment, orthostatic hypotension, delirium, mobility problems, and incontinence. Similarly, OHID identifies muscle weakness, poor balance, visual impairment, polypharmacy, and environmental hazards as falls risk factors. For the purposes of this study, we have also extracted dementia and frailty as risk factors.

To collate and evaluate concerns raised by coroners, we classified and identified repeated themes using directed content analysis [20]. This allowed us to highlight cases involving previously recognised concerns, and to explore concepts, drawing similarities and disparities from the data. Subgroup analyses were undertaken according to the location of fall.

To calculate response rates to PFDs, we used the 56-day legal requirement [11], to classify responses as ‘on time’, ‘late’, or ‘unspecified’ (where a date was not present). We calculated the average response rate and frequency for individuals and organisations. Responses from organisations in receipt of fall-related PFDs were synthesised and classified by type of change initiated using content analysis.

### Software

We used Datawrapper [21] to produce bar charts, pie charts and the choropleth map. We used SankeyMatic [22] to produce the Sankey diagram of fall location and death location. We used R (version 4.1.1), including the pdftools package, to create the web scraper that downloads and screens PFDs, which is openly available [15].

## Results

### Case characteristics

There were 527 fall-related PFDs and deaths between July 2013 and 16 November 2022 in England and Wales (12.5% of all PFDs). The rate of fall-related PFDs has persisted above 10% from 2013-2022 (Supplementary Appendix Figure 2, Supplementary Appendix Table 1). A median of 1.03% (IQR: 0.97-1.17) of all fall-related deaths in England and Wales, as per NHS Digital Statistics, were written into PFDs each year.

**Figure 2.**
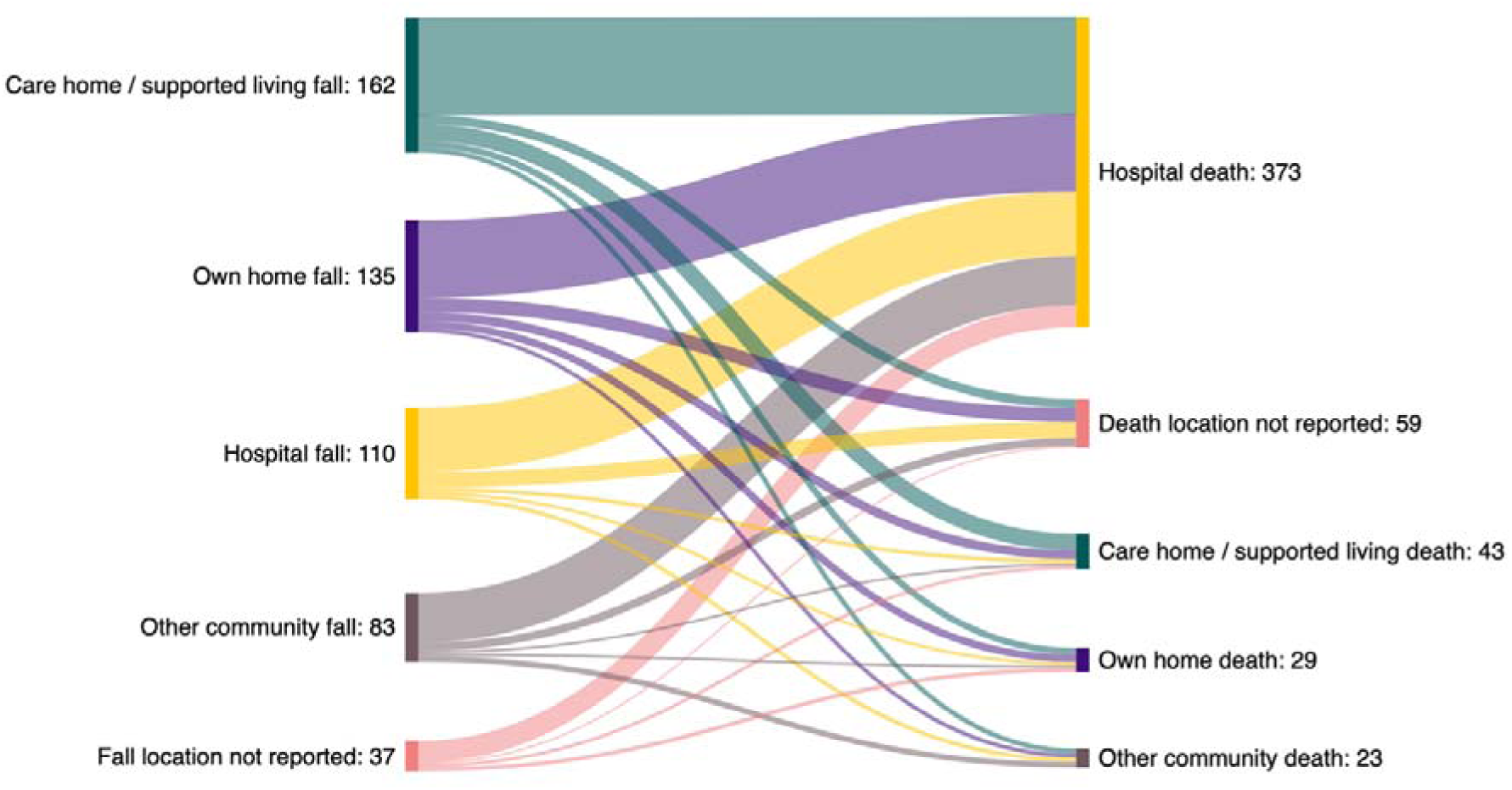
Sankey diagram comparing the location of initial fall and location of eventual death (n).

The median age at death was 82 years old (IQR: 70-89 years; n=340) (Figure 1), and 4,653 years of life were lost overall, with a median of 8 years lost per person (IQR: 5-17, n=340). Only 16 (3%) individuals were less than 30 years old at the time of death. Just over half (53.7%; n=283) of those who died were male.

The 527 PFDs were written across 80 coroner jurisdictions (Supplementary Appendix Table 3). Coroners in Northwest England wrote the most (26.6%) fall-related PFDs (Figure 3A, Supplementary Appendix Table 4), followed by South East England (14.4%), and Wales (10.1%). However, the highest number of fall-related PFDs as a proportion of the total PFDs from that region came from Wales (20.5%), North East England (19.9%), and North West England (16.5%) (Figure 3B, Supplementary Appendix Table 4).

**Figure 3.**
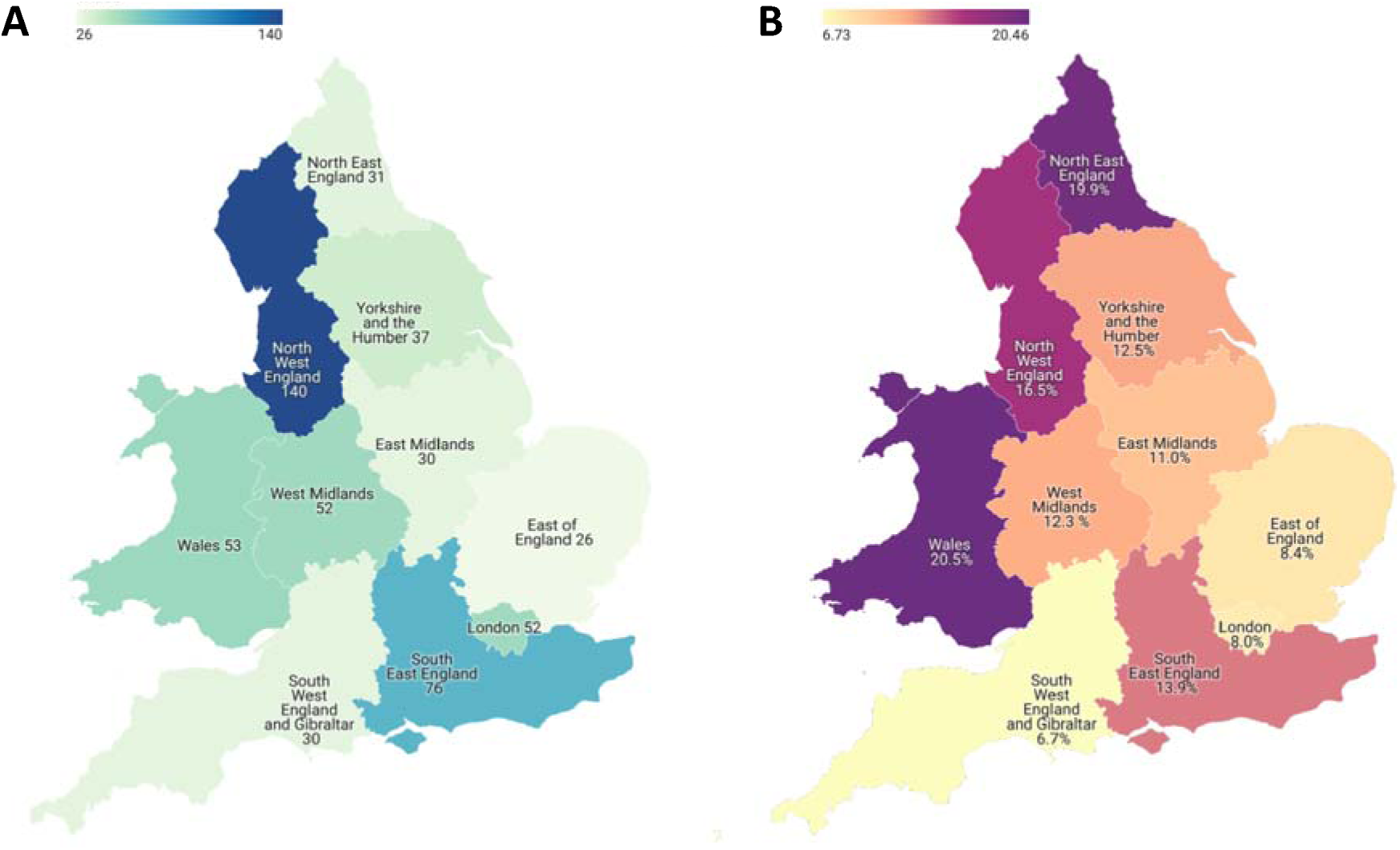
A) Raw number of fall-related PFDs published per region, 2013-2022. B) Number of fall-related PFDs published per region expressed as a percentage of all PFDs published by that region, 2013-2022.

On the Judiciary website, 56.4% of fall-related PFDs were classified under the “Hospital Deaths” report type, followed by “Care Home Health related deaths” (23.7%) and “Emergency Services related deaths” (12.3%) (Supplementary Appendix Table 5).

### Risk factors and causes of deaths

The most common risk factor identified was delirium/dementia (20.3%), followed by mobility problems (19.0%), and environmental hazards (15.4%) (Supplementary Appendix Table 6). 8.35% (n=44) of PFDs identified the individual as being frail. Out of the 111 falls that occurred in an inpatient environment, 68.7% did not describe any National Audit of Inpatient Falls (NAIF) risk factors (Supplementary Appendix Table 7). Following their fall, 51.6% of cases experienced a fracture, 35.9% had a major bleed, and 38.7% sustained a head injury (Supplementary Appendix Table 8).

After categorisation using ICD-11 codes, 34.0% of deaths were due to infection-related causes, 25.8% due to head injuries and neurovascular sequelae, and 11.5% due to cardiovascular causes (Supplementary Appendix Table 9). The most common specific causes of death were pneumonia (28.7%), traumatic subdural haematoma (17.8%), and pulmonary thromboembolism (4.6%). 11.4% of PFDs did not specify an explicit cause of death.

### Concerns raised by coroners

Coroners expressed 1,219 individual concerns in the 527 fall-related PFDs. Using content analysis, these concerns were categorised into 23 distinct themes within four higher-order chronological groups. These four groups were: before the fall, response time to the fall, medical care following the fall, and general concerns (Figure 4, Supplementary Appendix Table 10). 67.9% of concerns originated from falls in community settings, and 24.7% from falls in hospital (Supplementary Appendix Table S11).

**Figure 4.**
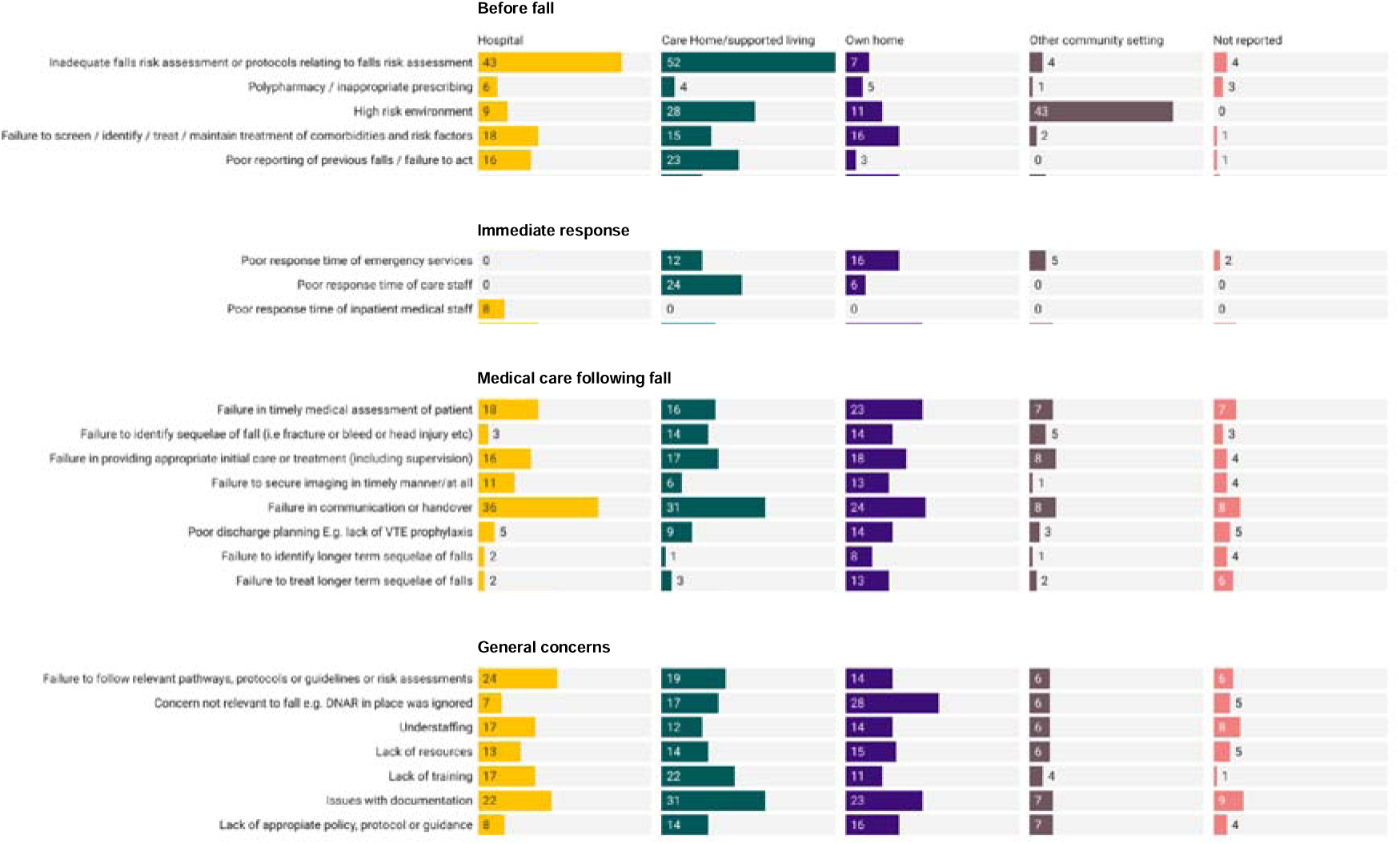
Coroner concern grouped by chronology of concern related to fall, and fall location

25.8% of all concerns were related to events before the fall. 20.9% of PFDs expressed concern surrounding inadequate falls risk assessments or protocols relating to falls risk, while 17.3% of PFDs expressed concern over high-risk environments (Supplementary Appendix Table 10). In the subgroup analysis by location of fall, concerns regarding falls risk assessments were raised in around a third of hospital (38.7%) and care home (32.3%) cases (Figure 4, Supplementary Appendix Table 10). Concerns around high risk environments were especially raised in care homes (17.4%) and other community settings (52.4%) (Figure 4, Supplementary Appendix Table 10).

5.9% of all concerns were related to the response time following the fall, most commonly poor response time of emergency services (6.6% of PFDs) (Supplementary Appendix Table 10). 49.3% of concerns expressed regarding poor immediate response time were in cases where falls occurred in care settings, and 30.1% in cases where falls happened in own homes (Figure 4, Supplementary Appendix Table 10).

37.8% of all concerns were related to events in the medical care following the fall, most commonly related to failures in communication or handover (20.3%) (Supplementary Appendix Table 10). 29.8% of all concerns were general concerns encompassing any time point around the fall. The most common general concern was regarding issues with documentation, which was reported in 17.5% of PFDs (Supplementary Appendix Table 10).

### Responses by organisations to fall-related PFDs

Coroners sent 730 PFDs to 35 categories of organisation (Supplementary Appendix Table 12) most commonly NHS organisations (49.5%). Overall, only 56.7% of reports had received a response at the time of data extraction, with just 37.7% of reports receiving a response within the 56 statutory days (Supplementary Appendix Table 12).

69.8% of responses acknowledged the issues raised by coroners and initiated new changes to address their concerns (Supplementary Appendix Table 13). This included 74.9% and 84.8% of responses from NHS organisations and care organisations, but only 45.6% and 48% responses from government and professional bodies respectively. 8.7% of responses did not acknowledge or agree with coroner concerns, including 22.2% of responses from ‘other’ organisations (Figure 5A, Supplementary Appendix Table 13).

**Figure 5.**
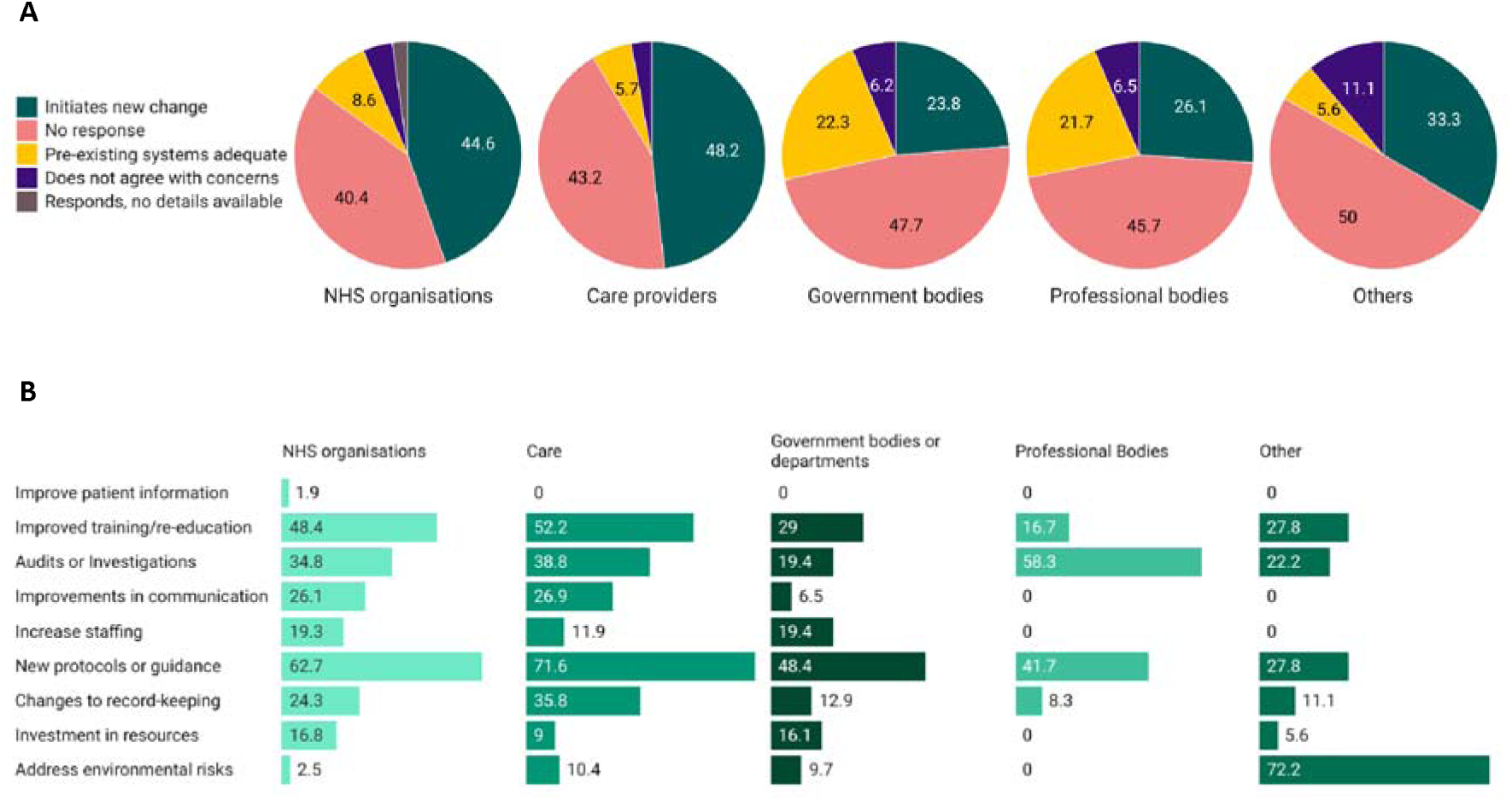
A) Initial higher-order response classification by organisation type. B) Classification of changes initiated by organisation (as a percentage) when made in response to coroner concerns.

Changes initiated by organisations to address coroner concerns were analysed and categorised into nine themes (Figure 5B, Supplementary Appendix Table 14). The most common actions including implementation of new protocols or guidance (58.5% of all responses), improved training or re-education (44.6%), and initiation of audits or investigations (34.3%). Notably, 72.2% of responses detailed changes initiated by ‘other’ organisations addressed environmental falls risk.

## Discussion

### Summary of findings

Between July 2013 and November 2022, there were 527 fall-related deaths in England and Wales where coroners determined that action should be taken to prevent future deaths. These cases predominantly occurred in older adults, who commonly fell in the community before dying in hospital. Around half of cases explicitly had at least one risk factor known to increase the risk of falls, and a high proportion of cases sustained a significant injury post-fall, including fractures, bleeds, and head injuries.

Coroners expressed a wide range of concerns spanning risk assessments before falls to issues with communication, documentation and care following the fall. These concerns were sent to hundreds of organisations, predominantly the NHS, care providers, and government bodies, but around half of reports have yet to receive a response. Organisations that responded commonly initiated audits to investigate events further, implemented new protocols related to falls or care, and sought to improve education and training.

### Comparison with existing literature

Several studies have investigated coroners’ concerns in PFDs [23-29], yet none have focussed on fall-related deaths. In a study investigating anticoagulant-using cardiovascular disease-related PFDs [24], concerns related to inadequate use of protocols were raised in 36.0% of cases, a concern similarly prevalent in our own fall-related cases. Similarly, coroners raised concerns around communication, seen in 20.3% of our cases, in 26.0% of medicines-related PFDs [23].

These studies have similarly found poor response rates to PFDs, a concern highlighted in the 2021 report by the House of Commons Justice Select Committee [30], and quantified in a recent quality analysis of all PFDs [31]. To our knowledge only one previous report has studied organisational responses to coroners concerns, in a limited number of medication-related deaths [32]. This study found that improving systems, performing audits, and implementing new policies were common responses, in line with our own fall-related findings. 16.4% of responses outlined no action or stated that current policies were sufficient, lower than the proportion among fall-related PFDs (28.3%).

Compared to the National Audit of Inpatient Falls (NAIF) [33], which examines inpatient falls in England and Wales where a fracture occurs, reporting of risk factors is far lower among PFDs. For example, mobility issues were cited in 92% of NAIF cases, but only 16.2% of PFD cases. This is likely to represent under-reporting of risk factors by coroners rather than an actual absence of risk factor among cases, despite identification of risk factors being an important aspect of falls prevention. Our included cases showed high rates of fractures post-fall, in line with data from the Global Burden of Disease Study 2017, which showed that fractures are the most common injury post-fall worldwide [34].

### Strengths and limitations

Our study forms part of the growing research that analyses samples and subgroups of PFDs [23-28], a data source that is used by the NHS Patient Safety Strategy [7] and begins to address several strategic research needs [9]. We used reproducible data collection methods to draw information from a large sample size of preventable death cases from both inpatient and community settings. We extracted information allowing comparison with national falls audits and describe organisational responses as well as concerns. There are some important limitations associated with using information from PFDs. Firstly, there is established inter-regional and inter-coroner variability in PFD writing [31], the latter being especially pertinent when considering the relative subjectivity of coroner concerns. Only around 1% of national fall-related deaths were written into a PFD each year, which is likely an underestimate of the true number of preventable deaths related to falls. Secondly, PFDs do not routinely include certain demographic data, such as ethnicity, gender, or socioeconomic status, and the reporting of age is inconsistent. Although we included deaths deemed to be preventable where a fall contributed to the death, in a few cases some preventable factors identified may have related to other fall-independent aspects of the patients’ healthcare. Finally, in instances where organisations have made changes to address coroners’ concerns, it is not possible to verify whether the changes have been implemented and whether the changes reduced falls and improved fall-related outcomes.

### Implications for practice

One in five people in the UK will be ≥65 years old by 2030 [35], a population in which 30% of people are falling at least once a year [36]. Falls are estimated to cost the NHS more than £2.3 billion per year [36]. Therefore, increased attention to falls prevention and falls management is urgently required to improve outcomes for patients at risk of falls. Indeed, supporting people to age well and improving support to those living in care homes was highlighted in The NHS Long Term Plan 2019 as a key priority [8]. Sharing lessons and actions taken following a PFD may help direct quality improvement efforts to reduce harm from falls.

Our study found that the majority of falls leading to preventable death happened in the community. The large NAIF audit exists to track the quality of prevention and care of inpatients sustaining a fall and fracture, but no comparable national audit exists for falls in the community. Our findings support an ongoing requirement for high quality, cost-effective and easily implementable falls risk assessment and prevention strategies for community health practitioners [37]. NICE supports the use of multifactorial interventions in the community [36], and a meta-analysis of 41 trials has shown that these may be successful in reducing the rate of falls [38]. There is concern however that the burden of prevention would fall to general practitioners (GPs), who may face multiple barriers to implementing the guidance outlined by NICE [39]. New risk prediction models based on large data sets may help identify patients who are most at risk and may benefit from early intervention [40].

Implementation of new protocols or guidance was the most common action taken by organisations in receipt of fall-related PFDs, but successful implementation of these changes is similarly important. Research demonstrates that success in reducing rates of falls in care homes is driven by effective staff-training to increase awareness of falls risk, local staff champions advocating an all-staff approach, and fostering attitudes that falls can be prevented [41]. A systematic review of falls prevention interventions in hospital inpatients found that patient and staff education in particular yielded significant reductions in falls rates [42], however previous reviews of guidelines have shown a relative paucity of education recommendations [43]. A separate review highlighted leadership support, engagement of all staff in falls prevention, and attitude change as further important aspects of prevention [44].

Not all falls are preventable, however morbidity can be significantly reduced by targeting coroner concerns regarding the post-fall period. Poor sharing of information, including verbal communication between staff and documentation, regarding patient care is a known cause of patient harm [45]. Further investment in communication channels in addition to staff training, including technology for documentation, may be required to minimise these harms.

In addition to the intrinsic health-based risk factors highlighted by NICE, our study showed a high-risk environment was cited by coroners in 17.3% of cases, especially within community settings. A prospective study highlighted the most common environmental factors in nursing homes that were significantly associated with occurrence of falls as inadequate or inappropriate handrails, unsafe ground, and insufficient lighting [46]. Ensuring safe healthcare environments remains a key potential area for quality improvement for healthcare providers both in hospitals and care homes, especially for vulnerable individuals at risk of falls.

### Conclusions

Fall-related PFDs provide valuable lessons in informing falls prevention and management in clinical practice. As the number of people ≥65 years old and at risk of falls in the population increases, implementation of their findings will improve patient safety and reduce morbidity and mortality from falls.

### Availability of data and materials

Protocols and study materials used for data synthesis are openly available on the Open Science Framework [13]. Demographic information from all PFD cases is openly available from the Preventable Deaths Tracker [16]. The code used to download and screen PFDs is openly available [15].

## Funding

No funding was obtained for this study. The National Institute for Health Research (NIHR) School for Primary Care Research (SCPR) provided funding to establish the Preventable Deaths Tracker website (2021-2022): https://preventabledeathstracker.net/

### Author contributions

FD conceptualised the project and wrote the code used to download and screen the PFDs. GCR created the Preventable Death Tracker and established the methodology to analyse PFDs. FD and CP wrote the study protocol. All authors were involved in screening the PFDs for inclusion. CP and KS performed the data extraction and data synthesis and wrote the manuscript. All authors were involved in interpretation of the results. All authors have read and endorse the findings of the final manuscript.

## Data Availability

All data produced in the present study are available upon reasonable request to the authors

