## Supplementary appendix for "Preventable deaths involving falls in England and Wales, 2013-2022: a systematic case series of coroners’ reports"

### **Supplementary Appendix Figure 1.** Flow diagram of inclusion and exclusion of cases downloaded from the Judiciary website.

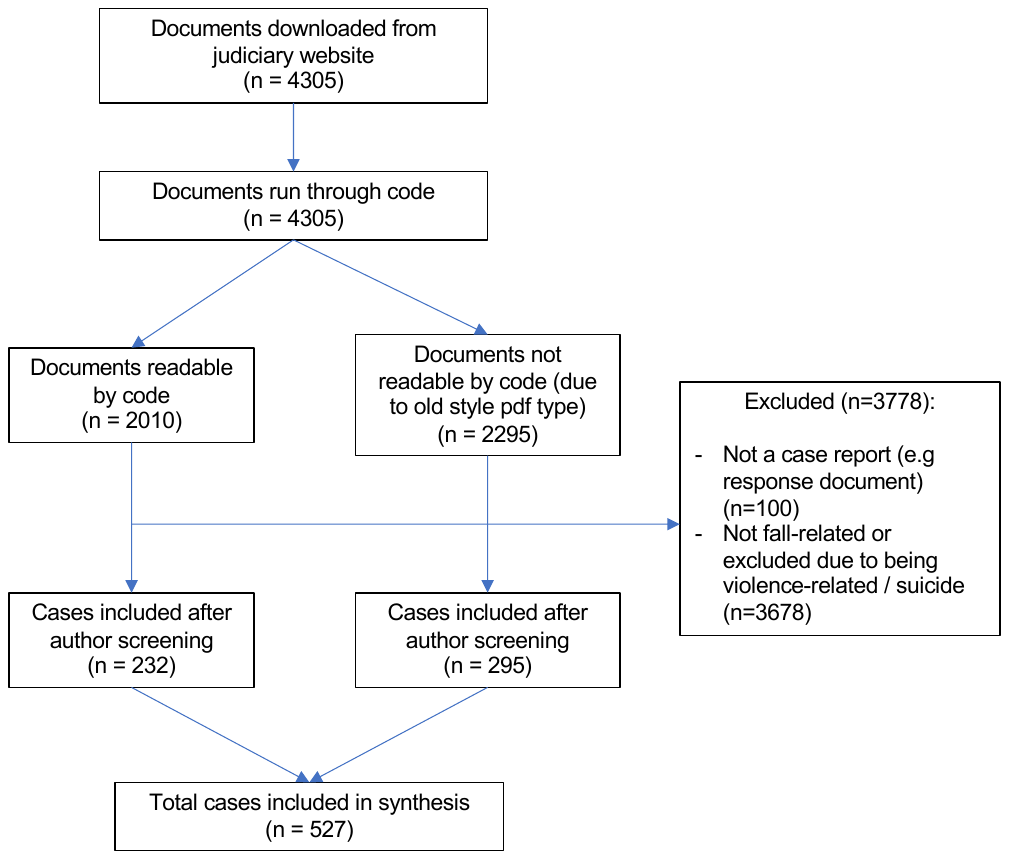

**Supplementary Appendix Figure 2.** The rate of fall-related Prevention of Future Deaths reports (PFDs) as a percentage of PFDs published yearly between 1 July 2013 and 16 November 2022 in England and Wales. Figure produced using Datawrapper.

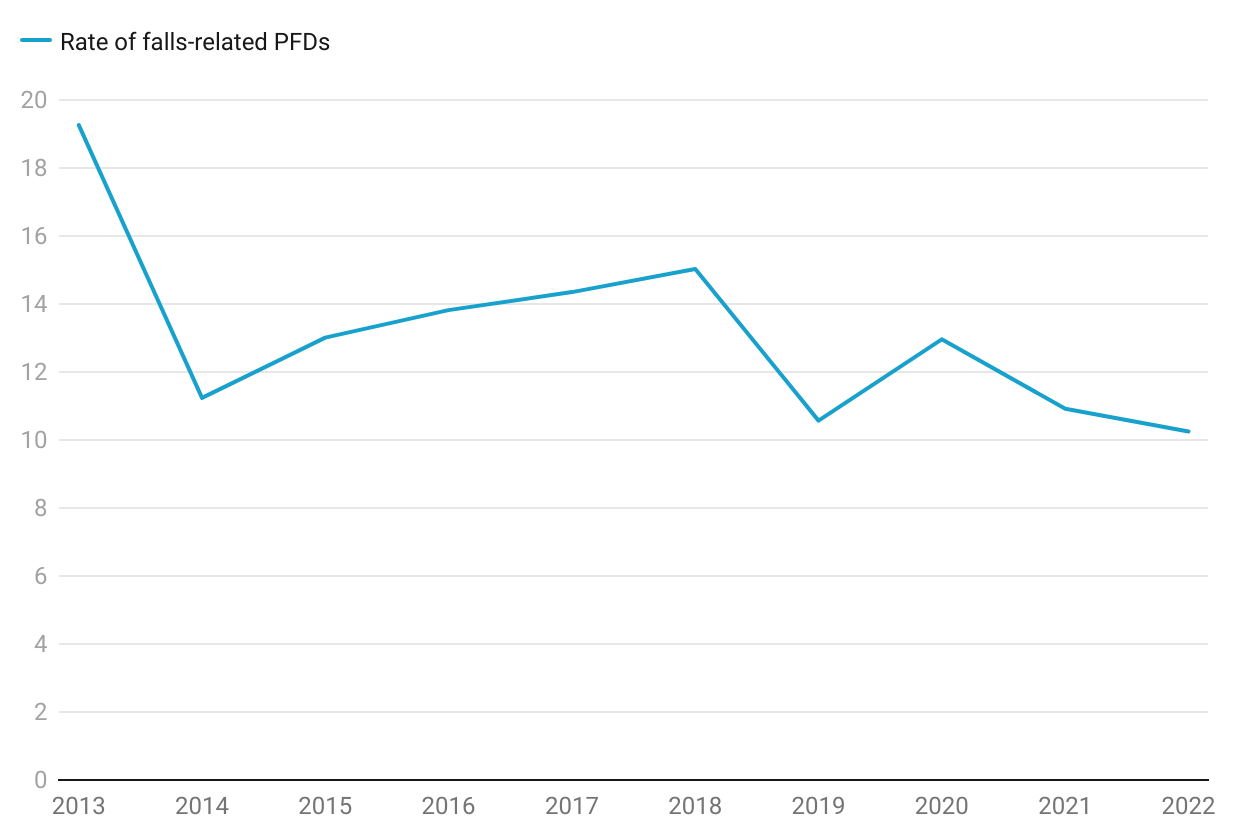

**Supplementary Appendix Table 1.** Number of falls-related PFDs by year, including as a percentage of all PFDs for that year and as a percentage of all falls-related deaths for that year as documented by the Office for National Statistics (ONS)

| **Year** | **Total number of PFDs** | **Number of falls-related PFDs** | **Rate of falls-related PFDs (%)** | **Number of ONS falls deaths** | **ONS falls deaths written into PFDs (%)** |
| --- | --- | --- | --- | --- | --- |
| 2013 | 135 | 26 | 19.26 | 4230 | N/A† |
| 2014 | 614 | 69 | 11.24 | 4610 | 1.50 |
| 2015 | 492 | 64 | 13.01 | 5415 | 1.18 |
| 2016 | 398 | 55 | 13.82 | 5600 | 0.98 |
| 2017 | 446 | 64 | 14.35 | 5535 | 1.16 |
| 2018 | 386 | 58 | 15.03 | 6085 | 0.95 |
| 2019 | 596 | 63 | 10.57 | 6100 | 1.03 |
| 2020 | 301 | 39 | 12.96 | 6410 | 0.61 |
| 2021 | 476 | 52 | 10.92 | N/A* | N/A* |
| 2022 | 361 | 37 | 10.25 | N/A* | N/A* |

*Falls deaths data not available for this year

†PFD publication commenced in July 2013, so not able to calculate rate for this year

**Supplementary Appendix Table 2.** Location of falls and deaths of the deceased as detailed in fall-related Prevent Future Death reports (PFDs).

| **Location** | **Percentage of falls, % (N)** | **Percentage of deaths, % (N)** |
| --- | --- | --- |
| Hospital | 21.1  (111) | 70.8  (373) |
| Care Home/Supported Living | 30.6  (161) | 8.2  (43) |
| Own Home | 25.8  (136) | 5.5  (29) |
| Other Community Setting | 15.6  (82) | 4.4  (23) |
| Not reported | 7.0  (37) | 11.1  (59) |

**Supplementary Appendix Table 3.** Classification of fall-related Prevention of Future Death reports (PFDs) according to coroner area described in the report.

| **Coroner Area** | **Number of PFDs (n=527)** |
| --- | --- |
| Avon | 8 |
| Bedfordshire & Luton | 9 |
| Berkshire | 1 |
| Birmingham and Solihull | 13 |
| Black Country | 19 |
| Blackburn, Hyndburn & Ribble Valley | 5 |
| Blackpool & Fylde | 7 |
| Brighton and Hove | 7 |
| Camarthenshire and Pembrokeshire | 2 |
| Cambridgeshire & Peterborough | 1 |
| Cardiff & the Vale of Glamorgan | 3 |
| Central and South East Kent | 5 |
| Cheshire | 1 |
| Cornwall & the Isle of Scilly | 7 |
| County Durham & Darlington | 12 |
| Coventry | 2 |
| Cumbria | 6 |
| Derby and Derbyshire | 9 |
| Dorset | 3 |
| East Riding and Kingston Upon Hull | 2 |
| Essex | 3 |
| Exeter & Greater Devon | 1 |
| Gloucestershire | 4 |
| Greater Manchester South | 14 |
| Gwent | 8 |
| Hampshire (Central) | 2 |
| Herefordshire | 3 |
| Isle of Wight | 1 |
| Lancashire and Blackburn with Darwen | 1 |
| Leicester City & South Leicestershire | 7 |
| Lincolnshire (Central) | 3 |
| Liverpool and Wirral | 5 |
| London (East) | 12 |
| London (North) | 2 |
| London (South) | 8 |
| London (West) | 2 |
| London Inner (North) | 19 |
| London Inner (South) | 5 |
| London Inner (West) | 4 |
| Manchester (City) | 7 |
| Manchester (North) | 9 |
| Manchester (South) | 69 |
| Manchester (West) | 14 |
| Mid Kent and Medway | 10 |
| Milton Keynes | 9 |
| Newcastle Upon Tyne and North Tyneside | 4 |
| Norfolk | 8 |
| North East Kent | 1 |
| Northamptonshire | 4 |
| North Northumberland and South Northumberland | 3 |
| North Wales (East & Central) | 15 |
| North West Wales | 2 |
| North Yorkshire (West) | 1 |
| Nottinghamshire | 8 |
| Oxfordshire | 4 |
| Powys, Bridgend & Glamorgan Valleys | 6 |
| Plymouth Torbay and South Devon | 5 |
| Portsmouth & South East Hampshire | 13 |
| Sefton, St Helens and Knowsley | 1 |
| Shropshire, Telford & Wrekin | 1 |
| South Wales Central | 16 |
| South Yorkshire (East) | 8 |
| South Yorkshire (West) | 6 |
| Staffordshire South | 8 |
| Stoke-on-Trent & North Staffordshire | 6 |
| Suffolk | 2 |
| Sunderland | 9 |
| Surrey | 12 |
| Swansea, Neath and Port Talbot | 1 |
| Teeside | 3 |
| Warwickshire | 1 |
| West Sussex | 11 |
| West Yorkshire (East) | 8 |
| West Yorkshire (West) | 10 |
| Wiltshire & Swindon | 2 |
| Worcestershire | 2 |
| York | 2 |

**Supplementary Appendix Table 4.** Classification of fall-related Prevention of Future Death reports (PFDs) according to administrative regions of England and Wales.

| **Region** | **Number of falls-related PFDs (n=527)** | **Distribution of PFDs across different regions** | **PFDs per region (n=4205)** | **Rate of falls PFDs by region** |
| --- | --- | --- | --- | --- |
| East Midlands | 30 | 5.69% | 272 | 11.03% |
| East of England | 26 | 4.93% | 310 | 8.39% |
| London | 52 | 9.87% | 649 | 8.01% |
| North East England | 31 | 5.88% | 156 | 19.87% |
| North West England | 140 | 26.57% | 848 | 16.51% |
| South East England | 76 | 14.42% | 547 | 13.89% |
| South West England and Gibraltar | 30 | 5.69% | 446 | 6.73% |
| Wales | 53 | 10.06% | 259 | 20.46% |
| West Midlands | 52 | 9.87% | 422 | 12.32% |
| Yorkshire and the Humber | 37 | 7.02% | 296 | 12.50% |

**Supplementary Appendix Table 5.** Classification of fall-related Prevention of Future Death reports (PFDs) as per their categories on the Judiciary Website.

| **Category** | **Number of falls related PFDs** | **Percentage of total falls-related PFDs** |
| --- | --- | --- |
| Hospital related deaths | 297 | 56.36% |
| Care Home Health related deaths | 125 | 23.72% |
| Emergency services related deaths | 65 | 12.33% |
| Community Healthcare | 59 | 11.20% |
| Other related deaths | 51 | 9.68% |
| Accident at Work and Health and Safety related deaths | 12 | 2.28% |
| Wales prevention of future deaths reports (2019 onwards) | 11 | 2.09% |
| Mental health related deaths | 7 | 1.33% |
| Alcohol, drug, and medication related deaths | 5 | 0.95% |
| Product related deaths | 5 | 0.95% |
| Child death | 2 | 0.38% |
| Railway related deaths | 2 | 0.38% |
| State Custody related deaths | 2 | 0.38% |

Note that PFDs may be tagged with multiple categories.

**Supplementary Appendix Table 6.** Prevalence of description of falls risk factors (as identified by National Audit of Inpatient Falls (NAIF) and Office for Health Improvement and Disparities (OHID)) and frailty across falls-related PFDs

| **Risk factor** | **Number of falls-related PFDs** | **Percentage of falls-related PFDs** |
| --- | --- | --- |
| Frailty | 44 | 8.35% |
| No NAIF or OHID risk factor mentioned | 275 | 52.2% |
| **National Audit for Inpatient Falls (NAIF)** | | |
| Delirium/Dementia | 107 | 20.30% |
| Mobility problems | 100 | 18.98% |
| Visual impairment | 14 | 2.66% |
| Orthostatic Hypotension | 6 | 1.14% |
| Incontinence | 5 | 0.95% |
| No NAIF risk factors mentioned | 337 | 63.9% |
| **Office for Health Improvement and Disparities (OHID)** | | |
| Environmental Hazards | 81 | 15.37% |
| Poor balance | 60 | 11.39% |
| Muscle weakness | 37 | 7.02% |
| Visual impairment | 14 | 2.66% |
| Polypharmacy | 4 | 0.76% |
| No OHID risk factors mentioned | 356 | 67.6% |

**Supplementary Appendix Table 7.** Prevalence of falls risk factors (as identified by National Audit of Inpatient Falls (NAIF) across falls-related PFDs where falls occurred in an inpatient setting (n=111).

| **Risk factor** | **Number of inpatient PFDs** | **Percentage of inpatient PFDs** |
| --- | --- | --- |
| Frailty* | 3 | 2.7% |
| Delirium/Dementia | 20 | 18.02% |
| Mobility problems | 18 | 16.21% |
| Visual impairment | 2 | 1.80% |
| Orthostatic Hypotension | 0 | 0% |
| Incontinence | 1 | 0.90% |
| No risk factor mentioned | 76 | 68.7% |

*****NB Frailty is not included in NAIF risk factors

**Supplementary Appendix Table 8.** Percentage of falls followed by fracture, bleed, or head injury as detailed in fall-related Prevent Future Death reports (PFDs).

| **Sequelae of Fall** | **Percentage of Cases, % (N)** |
| --- | --- |
| Fracture | 51.6  (272) |
| Bleed | 35.9  (189) |
| Head Injury | 38.7  (204) |

**Supplementary Appendix Table 9.** Causes of death within fall-related PFDs accounting for ≥0.5% of cases, by ICD-11 criteria. Note that coding of an individual death can involve multiple ICD-11 codes.

| **ICD-11 coding for cause of death** | **Count of Case**  **(N)** | **Percentage of Cases**  **(%)** | **Specific** |
| --- | --- | --- | --- |
| **Infection** | **179** | **34.0** | **N/A** |
| CA40 | 151 | 28.7 | Pneumonia |
| 1G41 or 1G40 | 20 | 3.8 | Sepsis (with or without septic shock) |
| RA01.0 | 5 | 0.9 | COVID-19 |
| GC08 | 3 | 0.6 | UTI |
| **Head injuries/**  **Neurovascular** | **136** | **25.8** | **N/A** |
| NA07.6 | 94 | 17.8 | Traumatic subdural haematoma |
| 8B00 | 18 | 3.4 | Intracerebral haemorrhage |
| NA07.Z | 14 | 2.7 | Unspecified intracranial injury |
| NA0Z | 7 | 1.3 | Injuries to the head, unspecified |
| 8B24 | 3 | 0.6 | Hypoxic-ischaemic encephalopathy |
| **Cardiovascular** | **61** | **11.5** | **N/A** |
| BB00 | 24 | 4.6 | Pulmonary thromboembolism |
| BD1Z | 16 | 3.0 | Heart failure |
| MC82.4 | 7 | 1.3 | Cardiopulmonary arrest |
| BA41 | 5 | 0.9 | Acute myocardial infarction |
| NF0A.1 | 5 | 0.9 | Fat embolism, traumatic, not otherwise specified |
| BA6Z | 4 | 0.8 | Ischaemic heart disease, unspecified |
| **Fractures** | **7** | **1.4** | **N/A** |
| NA22.Z | 4 | 0.8 | Fracture of cervical spine |
| NC72 | 3 | 0.6 | Fracture of femur |
| **Other** | **52** | **9.9** | **N/A** |
| ND37 | 17 | 3.2 | Multiple unspecified injuries |
| MG4A | 10 | 1.9 | Multiple organ failure |
| MG2A | 6 | 1.1 | Ageing associated decline in intrinsic capacity |
| CB41 | 5 | 0.9 | Respiratory failure |
| NF08.1 | 5 | 0.9 | Drowning |
| 6D8Z | 3 | 0.6 | Dementia |
| CB26 | 3 | 0.6 | Haemothorax |
| ME24.9 | 3 | 0.6 | GI bleeding |
| **No specified cause of death** | **60** | **11.4** | **N/A** |

Abbreviations: GI, gastrointestinal; UTI, urinary tract infection.

**Supplementary Appendix Table 10. Overall concerns and concerns by location of fall, raised by coroners grouped by higher-order theme following content analysis.**

Overall concerns by each higher-order theme are expressed as a percentage of the total number of fall-related PFDs (N=527).

Concerns by each higher-order theme in the subgroup analyses for location of fall are expressed as a percentage of the total number of cases where a fall-occurred in a location (e.g., Hospital, N=111).

*E.g., DNAR in place was ignored.

| **Chronology**  **(N of concerns)** | **Concerns** | **Percentage of Cases Overall, %**  **(N)** | **Percentage of Cases by Location of Fall, %**  **(N)** | | | | |
| --- | --- | --- | --- | --- | --- | --- | --- |
|  |  |  | **Hospital** | **Care Home/**  **Supported Living** | **Own home** | **Other Community Setting** | **Not Reported** |
| **Before fall**  **(315)** | Inadequate falls risk assessment or protocols relating to falls risk assessment | 20.9  (110) | 38.7  (43) | 32.3  (52) | 5.1  (7) | 4.9  (4) | 10.8  (4) |
|  | High risk environment | 17.3  (91) | 8.1  (9) | 17.4  (28) | 8.1  (11) | 52.4  (43) | 0.0  (0) |
|  | Failure to screen/identify/treat/maintain treatment of comorbidities and risk factors | 9.9  (52) | 16.2  (18) | 9.3  (15) | 11.8  (16) | 2.4  (2) | 2.7  (1) |
|  | Poor reporting/lack of action following previous falls | 8.2  (43) | 14.4  (16) | 14.3  (23) | 2.2  (3) | 0.0  (0) | 2.7  (1) |
|  | Polypharmacy/inappropriate prescribing | 3.6  (19) | 5.4  (6) | 2.5  (4) | 3.7  (5) | 1.2  (1) | 8.1  (3) |
| **Immediate response**  **(73)** | Poor response time of emergency services | 6.6  (35) | 0.0  (0) | 7.5  (12) | 11.8  (16) | 6.1  (5) | 5.4  (2) |
|  | Poor response time of care staff | 5.7  (30) | 0.0  (0) | 14.9  (24) | 4.4  (5) | 0.0  (0) | 0.0  (0) |
|  | Poor response time of inpatient medical staff | 1.5  (8) | 7.2  (8) | 0.0  (0) | 0.0  (0) | 0.0  (0) | 0.0  (0) |
| **Medical care following fall**  **(462)** | Failure in communication or handover | 20.3  (107) | 32.4  (36) | 19.3  (31) | 17.6  (24) | 9.8  (8) | 21.6  (8) |
|  | Failure in timely medical assessment | 13.5  (71) | 16.2  (18) | 9.9  (16) | 16.9  (23) | 8.5  (7) | 18.9  (7) |
|  | Failure to follow relevant pathways/protocols/guidelines | 13.1  (69) | 21.6  (24) | 11.8  (19) | 10.3  (14) | 7.3  (6) | 16.2  (6) |
|  | Failure in providing appropriate initial care/treatment | 12.0  (63) | 14.4  (16) | 10.6  (17) | 13.2  (18) | 9.8  (8) | 10.8  (4) |
|  | Failure to identify clinical sequelae of fall | 7.4  (39) | 2.7  (3) | 8.7  (14) | 10.3  (14) | 6.1  (5) | 8.1  (3) |
|  | Poor discharge planning | 6.8  (36) | 4.5  (11) | 5.6  (6) | 10.3  (13) | 3.7  (1) | 13.5  (4) |
|  | Failure to treat long term sequelae of falls | 4.9  (26) | 1.8  (2) | 1.9  (3) | 9.6  (13) | 2.4  (2) | 16.2  (6) |
|  | Failure to identify long term sequelae of falls | 3.0  (16) | 1.8  (2) | 0.6  (1) | 5.9  (8) | 1.2  (1) | 10.8  (4) |
| **General concerns**  **(369)** | Issues with documentation | 17.5  (92) | 19.8  (22) | 19.3  (31) | 16.9  (23) | 8.5  (7) | 24.3  (9) |
|  | Concern not relevant to fall* | 12.0  (63) | 6.3  (7) | 10.6  (17) | 20.6  (28) | 7.3  (6) | 13.5  (5) |
|  | Understaffing | 10.8  (57) | 15.3  (17) | 7.5  (12) | 10.3  (14) | 7.3  (6) | 21.6  (8) |
|  | Lack of training | 10.4  (55) | 15.3  (17) | 13.7  (22) | 8.1  (11) | 4.9  (4) | 2.7  (1) |
|  | Lack of resources | 10.1  (53) | 11.7  (13) | 8.7  (14) | 11.0  (15) | 7.3  (6) | 13.5  (1) |
|  | Lack of appropriate policy, protocol, or guidance | 9.3  (49) | 7.2  (8) | 8.7  (14) | 11.8  (16) | 8.5  (7) | 10.8  (4) |

**Supplementary Appendix Table 11. Coroners’ concerns expressed in fall-related Prevention of Future Death Reports (PFDs) by chronology of concern relative to the fall, stratified by location of fall.**

Concerns are expressed as a percentage of total number of concerns for a given chronology.

| **Chronology of Concern** | **Percentage of Concerns, %**  **(N)** | | | | |
| --- | --- | --- | --- | --- | --- |
|  | **Hospital** | **Care Home/**  **Supported Living** | **Own Home** | **Other Community Setting** | **Not reported** |
| **Before fall** | 29.2  (92) | 38.7  (122) | 13.3  (42) | 15.9  (50) | 2.9  (9) |
| **Immediate response** | 11.0  (8) | 49.3  (36) | 30.1  (22) | 8.9  (5) | 2.7  (2) |
| **Medical care following fall** | 25.3  (117) | 25.1  (116) | 30.5  (141) | 8.9  (41) | 10.2  (47) |
| **General concerns** | 22.8  (84) | 29.8  (110) | 29.0  (107) | 9.8  (36) | 8.7  (32) |
| **Total** | 24.7  (301) | 31.5  (384) | 25.6  (312) | 10.8  (132) | 7.4  (90) |

**Supplementary Appendix Table 12**. Organisations and individuals who received the 527 falls-related Prevention of Future Deaths reports (PFDs) in England and Wales published between July 2013 and November 2022 and their response rates according to Regulation 29 of the Coroners (Investigations) Regulations 2013. Similar individual organisations have been grouped together for clarity, e.g charities.

| **Organisations** | **Report received**  **(N)** | **Responses**  **(N)** | **Response rate**  **(%)** | **Classification of responses** | | | |
| --- | --- | --- | --- | --- | --- | --- | --- |
|  |  |  |  | **On time**∓  **(%)** | **Late**  **(%)** | **Unspecified response date**  **(%)** | **No response (%)** |
| **NHS organisations** | **361** | **215** | **59.6** | **41.3** | **15.0** | **3.3** | **40.4** |
| **CCGs** | 8 | 3 | 37.5 | 12.5 | 25.0 | 0.0 | 62.5 |
| **NHS England** | 24 | 18 | 75.0 | 25.0 | 50.0 | 0.0 | 25.0 |
| **Ambulance services** | 29 | 19 | 65.5 | 44.8 | 13.8 | 6.9 | 34.5 |
| **NHS trusts/hospitals** | 231 | 130 | 56.3 | 42.4 | 9.5 | 4.3 | 43.7 |
| **General Practitioners** | 8 | 5 | 62.5 | 50.0 | 12.5 | 0.0 | 37.5 |
| **Welsh Health Boards** | 33 | 18 | 54.5 | 30.3 | 24.2 | 0.0 | 45.5 |
| **Mental health clinics/trusts** | 5 | 3 | 60.0 | 40.0 | 20.0 | 0.0 | 40.0 |
| **Community health organisations*** | 1 | 0 | 0.0 | 0.0 | 0.0 | 0.0 | 100.0 |
| **NHS Health and Social Care Integrated/Partnership Trusts** | 22 | 19 | 86.4 | 68.2 | 18.2 | 0.0 | 13.6 |
| **Care** | **139** | **79** | **56.8** | **41.0** | **7.2** | **8.6** | **43.2** |
| **Care or nursing home** | 112 | 61 | 54.5 | 41.1 | 3.6 | 9.8 | 45.5 |
| **Care providers** | 22 | 15 | 68.2 | 40.9 | 22.7 | 4.5 | 31.8 |
| **Housing associations or supported accommodation** | 5 | 3 | 60.0 | 40.0 | 20.0 | 0.0 | 40.0 |
| **Government bodies or departments** | **130** | **68** | **52.3** | **25.4** | **25.4** | **1.5** | **47.7** |
| **Department of Health and Social Care** | 52 | 33 | 63.5 | 17.3 | 44.2 | 1.9 | 36.5 |
| **Local Councils** | 54 | 25 | 46.3 | 33.3 | 11.1 | 1.9 | 53.7 |
| **Welsh Government** | 10 | 2 | 20.0 | 20.0 | 0.0 | 0.0 | 80.0 |
| **Government transport agencies** | 4 | 2 | 50.0 | 50.0 | 0.0 | 0.0 | 50.0 |
| **Public Health England** | 2 | 2 | 100.0 | 50.0 | 50.0 | 0.0 | 0.0 |
| **Health and Safety Executive** | 5 | 3 | 60.0 | 20.0 | 40.0 | 0.0 | 40.0 |
| **Department for Digital Culture, Media, and Sport** | 1 | 1 | 100.0 | 0.0 | 100.0 | 0.0 | 0.0 |
| **Department for Community and Local Government** | 1 | 0 | 0.0 | 0.0 | 0.0 | 0.0 | 100.0 |
| **Department for Education** | 1 | 0 | 0.0 | 0.0 | 0.0 | 0.0 | 100.0 |
| **Professional Bodies** | **46** | **25** | **54.3** | **34.8** | **17.4** | **2.2** | **45.7** |
| **NICE** | 10 | 6 | 60.0 | 50.0 | 10.0 | 0.0 | 40.0 |
| **CQC** | 27 | 14 | 51.9 | 25.9 | 22.2 | 3.7 | 48.1 |
| **Royal Colleges** | 2 | 1 | 50.0 | 0.0 | 50.0 | 0.0 | 50.0 |
| **Nursing and Midwifery Council** | 2 | 1 | 50.0 | 50.0 | 0.0 | 0.0 | 50.0 |
| **MHRA** | 2 | 2 | 100.0 | 100.0 | 0.0 | 0.0 | 0.0 |
| **Other independent health inspectorates** | 3 | 1 | 33.3 | 33.3 | 0.0 | 0.0 | 66.7 |
| **Other** | **54** | **27** | **50.0** | **37.0** | **7.4** | **5.6** | **50.0** |
| **Chief Coroner** | 2 | 0 | 0.0 | 0.0 | 0.0 | 0.0 | 100.0 |
| **Police and Fire services** | 5 | 0 | 0.0 | 0.0 | 0.0 | 0.0 | 100.0 |
| **Private transport agencies** | 5 | 3 | 60.0 | 40.0 | 0.0 | 0.0 | 40.0 |
| **Charities** | 6 | 4 | 66.7 | 66.7 | 0.0 | 0.0 | 33.3 |
| **Construction companies/agencies** | 6 | 3 | 50.0 | 50.0 | 0.0 | 0.0 | 50.0 |
| **Private property owners** | 11 | 7 | 63.6 | 27.3 | 18.2 | 18.2 | 36.4 |
| **Prison and prison regulators** | 2 | 0 | 0.0 | 0.0 | 0.0 | 0.0 | 100.0 |
| **Medical equipment manufacturers** | 4 | 3 | 75.0 | 75.0 | 0.0 | 0.0 | 25.0 |
| **Miscellaneous** | 13 | 7 | 53.8 | 38.5 | 7.7 | 7.7 | 46.2 |
| **Total** | **730** | **414** | **56.7** | **37.7** | **14.9** | **4.1** | **43.3** |

∓Within statutory 56 days of date report is sent to organisation.

*For example, sexual health services.

Abbreviations: CCG, Clinical Commissioning Groups; CQC, Care Quality Commission; MHRA, Medicines and Healthcare products Regulatory Agency; NHS, National Health Service; NICE, National Institute for Health and Care Excellence.

**Supplementary Appendix Table 13.** Organisations and individuals who received the 527 falls-related Prevention of Future Deaths reports (PFDs) in England and Wales published between July 2013 and November 2022 and the nature of their response.

| **Organisations** | **Percentage of Responses**  **(%)** | | | |
| --- | --- | --- | --- | --- |
|  | **Acknowledges concern and initiates new change to address concern** | **Acknowledges concern but pre-existing solutions deemed adequate** | **Responds but does not acknowledge/agree with concern** | **Responds, no details available** |
| **NHS organisations** | **74.9** | **14.4** | **7.0** | **3.7** |
| **CCGs** | 66.7 | 33.3 | 0.0 | 0.0 |
| **NHS England** | 44.4 | 44.4 | 11.1 | 0.0 |
| **Ambulance services** | 57.9 | 21.1 | 10.5 | 10.5 |
| **NHS trusts/hospitals** | 81.55 | 8.5 | 6.6 | 2.3 |
| **General Practitioners** | 60.0 | 40.0 | 0.0 | 0.0 |
| **Welsh Health Boards** | 61.1 | 16.7 | 5.6 | 16.7 |
| **Mental health clinics/trusts** | 100.0 | 0.0 | 0.0 | 0.0 |
| **Community health organisations*** | N/A | N/A | N/A | N/A |
| **NHS Health and Social Care Integrated/Partnership Trusts** | 89.5 | 10.5 | 0.0 | 0.0 |
| **Care** | **84.8** | **10.1** | **5.1** | **0.0** |
| **Care or nursing home** | 85.2 | 69.8 | 4.9 | 0.0 |
| **Care providers** | 86.7 | 6.7 | 6.7 | 0.0 |
| **Housing associations or supported accommodation** | 66.7 | 33.3 | 0.0 | 0.0 |
| **Government bodies or departments** | **45.6** | **42.6** | **11.8** | **0.0** |
| **Department of Health and Social Care** | 36.4 | 60.6 | 3.0 | 0.0 |
| **Local Councils** | 68.0 | 12.0 | 20.0 | 0.0 |
| **Welsh Government** | 0.0 | 100.0 | 0.0 | 0.0 |
| **Government transport agencies** | 50.0 | 50.0 | 0.0 | 0.0 |
| **Public Health England** | 50.0 | 50.0 | 0.0 | 0.0 |
| **Health and Safety Executive** | 0.0 | 33.3 | 66.7 | 0.0 |
| **Department for Digital Culture, Media, and Sport** | 0.0 | 100.0 | 0.0 | 0.0 |
| **Department for Community and Local Government** | N/A | N/A | N/A | 0.0 |
| **Department for Education** | N/A | N/A | N/A | 0.0 |
| **Professional Bodies** | **48.0** | **40.0** | **12.0** | **0.0** |
| **NICE** | 16.7 | 66.7 | 16.7 | 0.0 |
| **CQC** | 64.3 | 35.7 | 0.0 | 0.0 |
| **Royal Colleges** | 100.0 | 0.0 | 0.0 | 0.0 |
| **Nursing and Midwifery Council** | 100.0 | 0.0 | 0.0 | 0.0 |
| **MHRA** | 0.0 | 50.0 | 50.0 | 0.0 |
| **Other independent health inspectorates** | 0.0 | 0.0 | 100.0 | 0.0 |
| **Other** | **66.7** | **11.1** | **22.2** | **0.0** |
| **Chief Coroner** | N/A | N/A | N/A | N/A |
| **Police and Fire services** | N/A | N/A | N/A | N/A |
| **Private transport agencies** | 33.3 | 66.7 | 0.0 | 0.0 |
| **Charities** | 75.0 | 25.0 | 0.0 | 0.0 |
| **Construction companies/agencies** | 0.0 | 0.0 | 100.0 | 0.0 |
| **Private property owners** | 100.0 | 0.0 | 0.0 | 0.0 |
| **Prison and prison regulators** | N/A | N/A | N/A | N/A |
| **Medical equipment manufacturers** | 0.0 | 100.0 | 100.0 | 0.0 |
| **Miscellaneous** | 100.0 | 0.0 | 0.0 | 0.0 |
| **Total** | **69.8** | **19.6** | **8.7** | **1.9** |

*For example, sexual health services.

Abbreviations: CCG, Clinical Commissioning Groups; CQC, Care Quality Commission; MHRA, Medicines and Healthcare products Regulatory Agency; NHS, National Health Service; NICE, National Institute for Health and Care Excellence.

**Supplementary Appendix Table 14. Changes initiated by organisations in receipt of a fall-related PFD in response to coroners’ concerns, classified by type of change.**

Type of change is expressed as a percentage of the total number of responses from each type of organisation that agreed with coroners’ concerns and initiated changes.

*For example, sexual health services.

Abbreviations: CCG, Clinical Commissioning Groups; CQC, Care Quality Commission; MHRA, Medicines and Healthcare products Regulatory Agency; NHS, National Health Service; NICE, National Institute for Health and Care Excellence.

| **Organisations** | **Percentage of Responses** | | | | | | | | |
| --- | --- | --- | --- | --- | --- | --- | --- | --- | --- |
|  | **Improve patient information** | **Improved training/re-education** | **Audits or Investigations** | **Improvements in communication** | **Increase staffing** | **New protocols or guidance** | **Changes to record-keeping** | **Investment in resources** | **Address environmental risks** |
| **NHS organisations** | **1.9** | **48.4** | **34.8** | **26.1** | **19.3** | **62.7** | **24.3** | **16.8** | **2.5** |
| **CCGs** | **0.0** | **0.0** | **0.0** | **0.0** | **0.0** | **50.0** | **0.0** | **50.0** | **0.0** |
| **NHS England** | **0.0** | **37.5** | **25.0** | **50.0** | **12.5** | **75.0** | **12.5** | **12.5** | **12.5** |
| **Ambulance services** | **9.1** | **54.5** | **18.2** | **9.1** | **72.7** | **54.5** | **9.1** | **45.5** | **0.0** |
| **NHS trusts/hospitals** | **1.9** | **49.1** | **34.0** | **30.2** | **14.2** | **58.5** | **30.2** | **14.2** | **2.8** |
| **General Practitioners** | **0.0** | **66.7** | **0.0** | **0.0** | **0.0** | **66.7** | **33.3** | **0.0** | **0.0** |
| **Welsh Health Boards** | **0.0** | **54.5** | **63.6** | **0.0** | **27.3** | **63.6** | **0.0** | **9.1** | **0.0** |
| **Mental health clinics/trusts** | **0.0** | **33.3** | **66.7** | **33.3** | **0.0** | **66.7** | **33.3** | **33.3** | **0.0** |
| **Community health organisations*** | **N/A** | **N/A** | **N/A** | **N/A** | **N/A** | **N/A** | **N/A** | **N/A** | **N/A** |
| **NHS Health and Social Care Integrated/Partnership Trusts** | **0.0** | **47.1** | **41.2** | **23.5** | **23.5** | **88.2** | **17.6** | **17.6** | **0.0** |
| **Care** | **0.0** | **52.2** | **38.8** | **26.9** | **11.9** | **71.6** | **35.8** | **9.0** | **10.4** |
| **Care or nursing home** | **0.0** | **46.2** | **40.4** | **34.6** | **15.4** | **73.1** | **36.5** | **11.5** | **11.5** |
| **Care providers** | **0.0** | **69.2** | **38.5** | **0.0** | **0.0** | **69.2** | **38.5** | **0.0** | **0.0** |
| **Housing associations or supported accommodation** | **0.0** | **100.0** | **0.0** | **0.0** | **0.0** | **50.0** | **0.0** | **0.0** | **50.0** |
| **Government bodies or departments** | **0.0** | **29.0** | **19.4** | **6.5** | **19.4** | **48.4** | **12.9** | **16.1** | **9.7** |
| **Department of Health and Social Care** | **0.0** | **16.7** | **25.0** | **0.0** | **33.3** | **25.0** | **16.7** | **33.3** | **0.0** |
| **Local Councils** | **0.0** | **41.2** | **17.6** | **11.8** | **11.8** | **64.7** | **11.8** | **5.9** | **11.8** |
| **Welsh Government** | **N/A** | **N/A** | **N/A** | **N/A** | **N/A** | **N/A** | **N/A** | **N/A** | **N/A** |
| **Government transport agencies** | **0.0** | **0.0** | **0.0** | **0.0** | **0.0** | **0.0** | **0.0** | **0.0** | **100.0** |
| **Public Health England** | **0.0** | **0.0** | **0.0** | **0.0** | **0.0** | **100.0** | **0.0** | **0.0** | **0.0** |
| **Health and Safety Executive** | **N/A** | **N/A** | **N/A** | **N/A** | **N/A** | **N/A** | **N/A** | **N/A** | **N/A** |
| **Department for Digital Culture, Media, and Sport** | **N/A** | **N/A** | **N/A** | **N/A** | **N/A** | **N/A** | **N/A** | **N/A** | **N/A** |
| **Department for Community and Local Government** | **N/A** | **N/A** | **N/A** | **N/A** | **N/A** | **N/A** | **N/A** | **N/A** | **N/A** |
| **Department for Education** | **N/A** | **N/A** | **N/A** | **N/A** | **N/A** | **N/A** | **N/A** | **N/A** | **N/A** |
| **Professional Bodies** | **0.0** | **16.7** | **58.3** | **0.0** | **0.0** | **41.7** | **8.3** | **0.0** | **0.0** |
| **NICE** | **0.0** | **0.0** | **0.0** | **0.0** | **0.0** | **100.0** | **0.0** | **0.0** | **0.0** |
| **CQC** | **0.0** | **11.1** | **77.8** | **0.0** | **0.0** | **33.3** | **11.1** | **0.0** | **0.0** |
| **Royal Colleges** | **0.0** | **0.0** | **0.0** | **0.0** | **0.0** | **100.0** | **0.0** | **0.0** | **0.0** |
| **Nursing and Midwifery Council** | **0.0** | **100.0** | **0.0** | **0.0** | **0.0** | **0.0** | **0.0** | **0.0** | **0.0** |
| **MHRA** | **N/A** | **N/A** | **N/A** | **N/A** | **N/A** | **N/A** | **N/A** | **N/A** | **N/A** |
| **Other independent health inspectorates** | **N/A** | **N/A** | **N/A** | **N/A** | **N/A** | **N/A** | **N/A** | **N/A** | **N/A** |
| **Other** | **0.0** | **27.8** | **22.2** | **0.0** | **0.0** | **27.8** | **11.1** | **5.6** | **72.2** |
| **Chief Coroner** | **N/A** | **N/A** | **N/A** | **N/A** | **N/A** | **N/A** | **N/A** | **N/A** | **N/A** |
| **Police and Fire services** | **N/A** | **N/A** | **N/A** | **N/A** | **N/A** | **N/A** | **N/A** | **N/A** | **N/A** |
| **Private transport agencies** | **0.0** | **0.0** | **0.0** | **0.0** | **0.0** | **100.0** | **0.0** | **0.0** | **100.0** |
| **Charities** | **0.0** | **0.0** | **66.7** | **0.0** | **0.0** | **0.0** | **0.0** | **0.0** | **33,3** |
| **Construction companies/agencies** | **N/A** | **N/A** | **N/A** | **N/A** | **N/A** | **N/A** | **N/A** | **N/A** | **N/A** |
| **Private property owners** | **0.0** | **42.9** | **14.3** | **0.0** | **0.0** | **42.9** | **0.0** | **14.3** | **85.7** |
| **Prison and prison regulators** | **N/A** | **N/A** | **N/A** | **N/A** | **N/A** | **N/A** | **N/A** | **N/A** | **N/A** |
| **Medical equipment manufacturers** | **N/A** | **N/A** | **N/A** | **N/A** | **N/A** | **N/A** | **N/A** | **N/A** | **N/A** |
| **Miscellaneous** | **0.0** | **28.6** | **14.3** | **0.0** | **0.0** | **14.3** | **28.6** | **0.0** | **71.4** |
| **Total** | **1.0** | **44.6** | **34.3** | **21.5** | **15.6** | **58.5** | **23.9** | **13.5** | **9.3** |
